# Two-Year Outcomes from the PRESERVE Trial: Durable Oncologic Control Following Focal Irreversible Electroporation Ablation for Intermediate-Risk Prostate Cancer

**DOI:** 10.64898/2026.05.08.26352470

**Authors:** Jonathan A. Coleman, Arvin K. George, the PRESERVE Trial Investigators, the Society of Urologic Oncology Clinical Trials Consortium (SUO-CTC)

## Abstract

The PRESERVE trial (NCT04972097) is a prospective, single-arm pivotal IDE study evaluating focal irreversible electroporation (IRE) using the NanoKnife System for intermediate-risk prostate cancer. Men with Gleason Grade Group 2–3 disease underwent focal IRE and were followed for durability of oncologic control and safety. At 24 months, 68 patients completed follow-up with no new treatment failures identified. PSA levels were below baseline in 97% of patients, and one clinically triggered biopsy was negative for cancer. No new device- or procedure-related adverse events occurred beyond 12 months. These findings demonstrate durable efficacy and sustained safety of focal IRE.

## Background

Focal irreversible electroporation (IRE) ablation is an established tissue-sparing treatment strategy for localized prostate cancer that exploits non-thermal cell death to ablate targeted lesions while preserving adjacent critical structures. The PRESERVE trial (NCT04972097) is a prospective, non-randomized, single-arm pivotal IDE study evaluating focal IRE using the NanoKnife System (AngioDynamics, Inc.) in 121 patients with intermediate-risk prostate cancer, conducted across 17 U.S. clinical centers in collaboration with the Society of Urologic Oncology Clinical Trials Consortium (SUO-CTC). Primary 12-month results demonstrated an 80% freedom-from-treatment-failure rate among protocol-biopsied patients and were published in European Urology.^1^ The present report describes 24-month follow-up outcomes, evaluating the durability of oncologic control and the extended safety profile.

## Methods

Eligible patients had biopsy-confirmed Gleason Grade Group 2–3 (Gleason score 3+4 or 4+3), clinical stage ≤T2c intermediate-risk prostate cancer and underwent focal IRE ablation using the NanoKnife System. Patients who were not treatment failures at 12 months from participating sites who provided consent for continued follow-up constituted the 24-month analysis-eligible cohort. Clinically triggered biopsy was performed as indicated based on PSA kinetics or imaging findings. Treatment failure was defined as consistent with the 12-month primary endpoint definition (any amount of in-field Gleason scores for gradable cancer). Adverse events were graded per CTCAE v5.0. Patients provided informed consent. The study protocol was approved by institutional review boards and conducted as per the International Conference on Harmonization’s Good Clinical Practice guidelines.

## Results

Of 121 originally enrolled patients, 72 were analysis-eligible for the 24-month assessment; 68 (94.4%) of these patients completed the 24-month visit, reflecting strong cohort retention. No new treatment failures were identified among analysis-eligible completers at 24 months. One clinically triggered biopsy was performed, which was negative for any cancer. All but two patients (66/68, 97%) had a PSA at 24 months that was less than their baseline PSA. No new device- or procedure-related adverse events were reported in the interval between the 12- and 24-month assessments. The cumulative procedure-related adverse event profile remained consistent with that reported at 12 months.

## Conclusions

Twenty-four-month follow-up from the PRESERVE pivotal trial demonstrates durable oncologic control following focal IRE ablation for intermediate-risk prostate cancer, with no new treatment failures and a stable safety profile. These findings support the sustained efficacy of focal IRE as a tissue-sparing treatment option for appropriately selected patients and provide a robust, prospectively collected U.S. evidence base relevant to clinical guideline development and coverage policy deliberation. These results also correlate with findings from the Australian long-term (median 5 years) follow up experience.^2^

## Supporting information

List of the IRBs

## Data Availability

All data produced in the present work are contained in the manuscript.

## Trial Registration

ClinicalTrials.gov NCT04972097

