## Supplementary material for "Two-Year Outcomes from the PRESERVE Trial: Durable Oncologic Control Following Focal Irreversible Electroporation Ablation for Intermediate-Risk Prostate Cancer": List of the IRBs

### 16.1.3 List of IECs or IRBs, Written Information for Patient, and Sample Consent Forms

| Site No. and Name | Principal Investigator | Investigator's IRB | IRB Chair |
|---|---|---|---|
| 101 – University of Colorado Urology Clinic | Al Barqawi, MD | WCG IRB<br>1019 39th Ave SE, Suite 120<br>Puyallup, WA 98374 | |
| 102 – Memorial Sloan Kettering Cancer Center | Jonathan Coleman, MD | Memorial Sloan Kettering Cancer Center IRB<br>1275 York Avenue<br>New York, NY 10065 | R. Michael Tuttle, MD |
| 103 – Fox Chase Cancer Center | Andres Correa, MD | WCG IRB<br>1019 39th Ave SE, Suite 120<br>Puyallup, WA 98374 | |
| 104 – University of Florida Health | Wayne Brisbane, MD / Padraic O'Malley, MD | WCG IRB<br>1019 39th Ave SE, Suite 120<br>Puyallup, WA 98374 | |
| 105 – University of Texas Southwestern Medical Center | Jeffrey Gahan, MD | University of Texas Southwestern Medical Center IRB<br>5323 Harry Hines Blvd.<br>Dallas, TX 75390 | Ahamed Idris |
| 107 – Northshore University Health System | Brian Helfand, MD | WCG IRB<br>1019 39th Ave SE, Suite 120<br>Puyallup, WA 98374 | |
| 108 – USC Keck School of Medicine | Amir Lebastchi, MD | WCG IRB<br>1019 39th Ave SE, Suite 120<br>Puyallup, WA 98374 | |
| 109 – Mayo Clinic | Derek Lomas, MD | 200 First St SW<br>Rochester, MN 55905 | Taimur Sher, MD |
| 110 – Weill Cornell Medicine | Timothy McClure, MD | BRANY IRB<br>1981 Marcus Avenue, Suite 210<br>Lake Success, NY 11042 | Not published |
| 111 – Duly Health and Care | Ranko Miocinovic, MD | WCG IRB<br>1019 39th Ave SE, Suite 120<br>Puyallup, WA 98374 | |
| 112 – Duke Cancer Center | Thomas Polascik, MD | Duke University Health System IRB<br>Suite 900 Erwin Square<br>2200 West Main Street<br>Campus Box # 104026<br>Durham, NC 27705 | Dara Barnard, PharmD; Austin Joseph, JD, LLM; Louis F. Diehl, MD; Mark P. Donahue MD; |

| Site No. and Name | Principal Investigator | Investigator's IRB | IRB Chair |
|---|---|---|---|
| | | | Moria J. Smoski, PhD; Richard Lee, MD, MPH; Sharon L. Ellison, PharmD; Walter T. Lee, MD: Wanda Lakey, MD, MHS |
| 113 – Northwell Health – Lenox Hill Hospital | Art Rastinehad, MD | WCG IRB<br>1019 39th Ave SE, Suite 120<br>Puyallup, WA 98374 | |
| 114 – University of Cincinnati Medical Center | Abhinav Sidana, MD / Nilesh Patil, MD | WCG IRB<br>1019 39th Ave SE, Suite 120<br>Puyallup, WA 98374 | |
| 116 – University of California Irvine Medical Center | Edward Uchio, MD | WCG IRB<br>1019 39th Ave SE, Suite 120<br>Puyallup, WA 98374 | |
| 117 – Rush University Medical Center | Srinivas Vourganti, MD | Rush University Medical Center IRB<br>600 S. Paulina St.<br>Chicago, IL 60612 | John Cobb |
| 118 – New York University Urology Associates | James Wysock, MD | NYU Langone Health IRB<br>1 Park Avenue, 6th Floor<br>New York, NY 10016 | Stuart Katz, MD, PhD |
| 119 – Moffitt Cancer Center | Alice Yu, MD | Advarra IRB<br>6100 Merriweather Dr., Suite 600<br>Columbia, MD 21044 | Diego Astein, MD; Robert Blum, PharmDSusan Ebert, Luke Gelinas, Ran Goldman, MD; Amanda Highly, David Hiller; Dena Johnson; Frederick Kopec, JD; Daniel Kronish; MD: Christopher Martin, PharmD; Erin Odor; Cheri Pettey; Gail Povar, MD; Mary Ruwart; Sukhbir Singh, MD; Christian Westby |
| 121 – Atrium Health - Wake Forest Baptist Urology | Matvey Tsivian, MD | Wake Forest School of Medicine IRB<br>8th Floor, Hanes Building<br>1 Medical Center Boulevard<br>Winston-Salem, NC 27157 | Douglas Yoder |
| 122 – University of Rochester Medical Center | Thomas Frye, MD | WCG IRB<br>1019 39th Ave SE, Suite 120<br>Puyallup, WA 98374 | |

| Site No. and Name | Principal Investigator | Investigator’s IRB | IRB Chair |
|---|---|---|---|
| 124 – Veterans Affairs Ann Arbor Healthcare System | Arvin George, MD / Kristian Stensland, MD | VAAAHS IRB<br>U.S. Department of Veterans Affairs<br>810 Vermont Avenue, NW Washington DC 20420 | Michael M. Wang, MD |

**Subject Information**
For participation in the study:

| | |
|---|---|
| **TITLE:** | Pivotal Study of the NanoKnife System for Ablation of Prostate Tissue in an Intermediate Risk Patient Population |
| **PROTOCOL NO.:** | 2021-ONC-01<br>WCG IRB Protocol #20213709 |
| **SPONSOR:** | AngioDynamics |
| **INVESTIGATOR:** | Name<br>Address<br>City, State Zip<br>Country |
| **STUDY-RELATED PHONE NUMBER(S):** | [24-hour number is required] |

### 1 What is the purpose of this form?

You have been asked to take part in a research study. Studies are needed to learn more about new treatments, such as how well they work and how safe they are. Global laws require that all new medical devices being made undergo clinical studies that will provide more insight into how they work in patients to treat a disease.

Taking part in this study is voluntary. You do not have to take part in this study if you do not want to. Your decision to take part or not take part will in no way affect your current or future treatment.

Please read the following information carefully. If you agree to take part in this study, you will be asked to sign the attached consent forms. The study doctor and/or a member(s) of the study staff can answer any questions you may have and provide explanation. If you would like to take part in this study, please do not sign the consent forms until you understand the goals and risks of the study and the procedures involved and you have been informed of your rights. We thank you for your time and attention.

If you decide to take part in the study, a signed and dated copy of this Subject Information form and the Informed Consent Form will be given to you.

### 2 Why is this study being done?

This study is being run for research purposes. The study device, NanoKnife® System, is being tested for the treatment of early-stage prostate cancer. The purpose of this study is to learn more about the use of the NanoKnife System for the treatment of early-stage prostate cancer. The NanoKnife is a type of irreversible electroporation (IRE) device. "Electroporation" is a cellular destruction technology that can be used to treat cancer. It uses electrical pulses to make small holes in cells that result in cell death.

The use of this device for the treatment of prostate cancer is not approved or cleared by the Food and Drug Administration (FDA), which means the use of the device in this study is investigational. The safety and effectiveness of this device for the treatment of prostate cancer has not been established. Please be advised that the treatments for this study are for investigational purposes only.

All subjects in this study will undergo treatment with the NanoKnife system and then receive “standard of care” for the further treatment of prostate cancer. The standard of care for further treatment of prostate cancer will be determined by the hospital where you receive your treatment.

The study will be conducted by an investigator (study doctor) who is responsible for the study at your study site.

The sponsor of this study is AngioDynamics. AngioDynamics will also be responsible for running the study. The principal investigator, who is the person in charge of ensuring the ethical conduct of this study, is.

The investigator is also called the study doctor.

The study will be done at up to up to 20 sites in the United States. The name and address of your study site and the study doctor for your study site are listed in **Section 16** of this form.

Your taking part in the study is expected to last for 12 months. This includes follow-up checks. During this time, you will receive treatment with the NanoKnife **and then** standard of care treatment for prostate cancer. Up to 118 participants will take part in this study in the US.

The sponsor will have access to your medical records for up to four years after your participation in the study is complete, in order to continue to study the long-term effects of the NanoKnife treatment.

### 3 Why have I been invited to take part in this study?

You are being asked to take part in this study because your medical records show you would be a suitable candidate. You have been diagnosed with prostate cancer and you are a candidate for electroporation treatment.

### 4 What will happen during this study?

Before joining the study, you will be asked to tell the study doctor about any medication that you are taking.

You should tell your regular doctor or health care provider that you are taking part in this study. Any treatment or medication prescribed to you during this study must be brought to the attention of the study staff.

In this study, blood and urine samples will be collected for safety tests. During the study, the amount of blood drawn will be within the standard accepted guidelines for research purposes. In addition, an MRI and/or ultrasound exam will be performed at some visits, and you will undergo a follow-up prostate biopsy at Visit 8 (twelve months), or if a recurrence of your cancer is suspected.

The following tests and procedures will be performed while you are in the study:

- Visit 1 (Screening): Give consent to take part in the study; review of study entry rules; review medical history and demographic information such as date of birth, gender, and race. As part of your diagnosis of prostate cancer, you will have undergone an ultrasound and MRI assessment, had a biopsy of your prostate, and have had a blood test for Prostate-Specific Antigen (PSA).
- Visit 2 (Baseline): Physical exam including vital signs, height, and weight; blood and urine samples; and a review of current medications or treatments. You will be asked to complete four (4) distinct questionnaires in order to catalog your current health status. You will be given instructions on food, liquid and medication intake the day before the procedure.
- Visit 3 (Day of Procedure): You will be admitted to the hospital. You will receive an abbreviated physical exam, have a review of your medications or treatments, and have an enema to cleanse the lower portion of your bowel as required. After general anesthesia is administered, a catheter will be inserted through your penis into your bladder to drain urine during the NanoKnife procedure. The NanoKnife treatment, which is performed under ultrasound guidance, involves the placement of special needles into the prostate through the perineum, the region between your anus and scrotum. The ultrasound will confirm that the needles are in the correct position within the prostate, surrounding the prostate cancer. The study doctor will then use the NanoKnife System to generate pulses of electricity between the needles, creating tiny holes in the prostate cells, causing them to die. Once the desired set of pulses is given between each needle pair, the needles will be removed and you will be brought to the recovery unit. Following your procedure, your medications will be reviewed and you will provide another urine sample if an infection is suspected. You may be required to stay in the hospital overnight. Some patients are discharged from the hospital with the catheter still in place. If this occurs, you will be instructed on how to care for the catheter. All subjects will be prescribed a course of antibiotics following the procedure. Those discharged with the catheter still inserted will be prescribed additional antibiotics.
  - Post-Procedure (Day 3-10): 3-10 days after the procedure, you come in for a check up to make sure you are doing as expected after your treatment. If your study doctor has any concerns, you may have a follow-up MRI. You will provide a urine sample if an infection is suspected. If an MRI and a urine sample are not required, this visit may be conducted over the phone.
- Visit 4 (Day 30 – one month): Physical exam including vital signs and weight; blood (including a PSA check) and urine (if an infection is suspected) samples; review of current medications and treatments. You will be asked to complete the four (4) distinct questionnaires to catalog your current health status.
  - Visit 5 (Day 90 – three months): Physical exam including vital signs and weight; blood (including a PSA check) and urine (if an infection is suspected) samples; review of current medications and treatments. You will have a follow-up MRI to confirm that the NanoKnife treatment was completed as intended and to check for any

cancer recurrence or progression. If your MRI results show that cancer remains or there is new cancer, you will undergo an early follow-up biopsy of your prostate. If this biopsy is positive, your study doctor will discuss your treatment options with you. You will be asked to complete the four (4) distinct questionnaires to catalog your current health status.

- Visit 6 (Day 180 – six months): Physical exam including vital signs and weight; blood (including a PSA check) and urine (if an infection is suspected) samples; review of current medications and treatments. You will be asked to complete the four (4) distinct questionnaires to catalog your current health status.
- Visit 7 (Day 270 – nine months): Physical exam including vital signs and weight; blood (including a PSA check) and urine (if an infection is suspected) samples; review of current medications and treatments. You will be asked to complete the four (4) distinct questionnaires to catalog your current health status.
- Visit 8 (Day 365 – twelve months): Physical exam including vital signs and weight; blood (including a PSA check) and urine (if an infection is suspected) samples; review of current medications and treatments. You will undergo an MRI and a biopsy of your prostate at this time. You will be asked to complete the four (4) distinct questionnaires to catalog your current health status.

### 5 What are the risks of taking part in this study?

You may have some unwanted effects and symptoms as a result of electroporation treatment with the NanoKnife System. The significant risks associated with electroporation treatment and the NanoKnife System may include:

- Cardiac Arrhythmia (improper heart beating)
- Muscle Injury/Muscle Weakness
- Bowel Injury which could necessitate surgical repair
- Nerve damage that could lead to loss of feeling in the penis and/or erectile dysfunction (“impotence”)
- Need for surgical intervention/repair
- Urinary incontinence
- Urinary retention
- Bleeding following probe placement which could lead to transfusions
- Infection
- Fever
- Pain
- Bruising (hematoma)
- Coagulopathy
- Abscess
- Hemolysis (destruction of red blood cells)
- Hyperbilirubinemia (breakdown of aged blood cells)
- Renal Dysfunction/Renal Toxicity (kidney failure)
- Liver Dysfunction (liver failure)
- Pseudoaneursym
- Thrombosis
- Fistula formation
- Injury to adjacent structures (nerve, vessel, duct. tissues)
- Cancer spread*

- Deep Vein Thrombosis (blood clot in the deep vein)
- Microemboli and Thrombi (blood clot)
- Death

*It is unclear whether your cancer material released from the IRE cells can be potentially absorbed by healthy cells, potentially leading to spread of your cancer, which might not be found for several months.

**Risks of Anesthesia**
The common side effects of general anesthesia include nausea and vomiting, dry mouth, sore throat or hoarseness, chills, confusion, dizziness, muscle aches, itching and difficulty urinating. Serious, life threatening side effects such as heart rhythm disturbances, strokes or accidents causing brain damage can occur. During your surgical procedure, while you are under general anesthesia, you will receive a drug (a paralytic agent) to control possible muscle contractions that may occur when electrical impulses are applied at or near muscles. These agents may have serious side effects and require monitoring by an anesthesiologist.

**Risks of Post-Operative Biopsy**
The post-operative biopsy will require ultrasound visualization and guidance; risks associated with ultrasound are found below. Risks associated with the biopsy procedure may include pain, bleeding (hematoma), infection, damage to unintended structures (nerves, vessels, ducts, and tissues), and the potential of tumor seeding (cancer spread).

**Risks of Post-Operative Ultrasound Assessment**
Ultrasound assessments will be conducted through use of an ultrasound probe placed in the rectum. This will require bowel prep prior to the procedure. It is possible that the ultrasound probe could damage the rectum or bowel or anal sphincter.

**Risks of Post-Operative MRI Assessment**
You may have risks from an MRI assessment if you have metallic objects in your body. You may also become anxious from lying in a tight space without moving.

**Risks of Antibiotics**
Antibiotics can cause nausea, vomiting, bloating and diarrhea. Allergic reactions can include coughing, wheezing and tightness in the throat.

**Other Risks**
Due to the investigational nature of this study, there may be other risks that are not yet known. If you have any unwanted effects, please make a note of them and report them to the study doctor at your next scheduled visit. **Please inform the study doctor immediately at the telephone number listed in Section 16 of any serious unwanted effects.**

Some procedures may cause you some pain or discomfort. For example, pain and/or bruising at the puncture site, bleeding and infection may occur during or after a blood draw. Swelling of a vein or in very rare cases, a blood clot, cannot be ruled out entirely.

It cannot be ruled out that your ability to operate machines or drive a car could be affected while taking part in this study.

### 6 Does taking part in this study provide any benefit?

Taking part in this study does not mean that you will benefit from the treatment you are given. The purpose of this study is to learn more about the study device.

While it is possible that treatment with the NanoKnife may improve your symptoms, you might not have any benefits from taking part in this study.

Although you may not receive any benefit yourself, your taking part in this study may provide new information about the treatment of prostate cancer.

### 7 What other treatments are available?

There are other drugs and procedures available to treat prostate cancer. Your study doctor will discuss these options with you.

You do not have to take part in this study to receive treatment for your cancer. If you do not take part in this study, you may have other treatment options such as those listed below:

- Getting treatment or care for your cancer without being in a study
- Taking part in another study
- Getting no treatment

For details on the benefits and risks of other treatments, please refer to the related product information or ask your study doctor.

### 8 What about confidentiality?

By signing this form, you agree to let the study doctor to allow certain groups to review information about your taking part in the study and your medical records. These might include the study sponsor, its designees, ethics committees/institutional review boards, and regulatory authorities such as the FDA. For more information on how your personal information is handled, please see the section titled, “Information Concerning Data Protection” at the end of this form.

All data and medical records tied to your taking part in this study will be kept private, except where required by law. If information about the study is published, it will be written in such a way that you cannot be recognized. You will be identified by a unique study number and not by your name whenever possible.

Data collected about you for the study may be included in reports. These reports will be submitted to authorities in various countries so that perhaps one day, the study device may be available to all patients who have prostate cancer and require this type of treatment.

During the study, a “monitor” will visit the study site. The monitor is responsible for making sure that the study is being performed properly. The monitor will have access to your original medical records (including confidential data that identifies you by name). The monitor will compare these records to study forms for correctness.

### 9 What will happen to my study samples?

Study staff will collect blood and urine samples from you as described in this form. Samples will be kept by a local laboratory and destroyed after testing.

Additionally, you will undergo a biopsy of your prostate at the 12-month visit, or sooner if a recurrence of your cancer is suspected. These biopsy samples may be stored and used for genomic testing.

By signing this informed consent form, you allow the study staff to provide these samples to the sponsor and others working with the sponsor. Your blood samples will be used to test for safety reasons.

### 10 Will it cost me anything to be in this study?

All costs for the required study visits, examinations, and laboratory procedures that are not part of routine medical care will be paid by AngioDynamics.

You or your insurance will be responsible for paying for the cost of any routine medical care that you would receive whether you participate in this study or not, unless you are told that such item or services will be supplied at no cost. If you have health insurance, the insurance may or may not pay for the costs associated with your participation in this study. You will have to pay for any co-payments, deductibles or co-insurance amounts that your insurance coverage requires. Before you decided to be in this study, you should contact your insurance provider to verify coverage.

You will receive fair compensation for your travel expenses to and from the study site. Please discuss compensation with the study staff.

### 11 What happens if something goes wrong?

Contact the study doctor if you feel that you have been injured or made sick because of being in this study **(see Section 16)**. Medical care will be provided.

If you suffer an injury which, in the reasonable judgment of Institution, was directly caused by the Study Device or any properly performed procedures required by the Protocol, Sponsor shall reimburse for the reasonable and necessary costs of diagnosis and treatment of any Study subject injury, including hospitalization, but only to the extent such expenses are not attributable to:

- Institution's negligence or willful misconduct;
- failure of Institution, the investigator, or any other Study personnel to adhere to the terms of the Study protocol or any written instructions (including, without limitation, instructions for use and operators manual, where appropriate) relative to the use of any product(s) used in the performance of the Study, or comply with applicable FDA or other government requirements, except to protect the safety and welfare of the Study subject;
- the natural progression of an underlying or pre-existing condition or events;

- the Study subject's failure to comply with instructions contained in the informed consent form executed by such Study subject or communicated to the Study subject by Study personnel; or
- a known risk of the Study device of the type being studied.

If you are injured as a result of being in this study you do not give up your right to pursue a claim through the legal system.

### 12 Do I have to take part in this study?

Taking part in this study is voluntary. You are free to refuse to take part or withdraw from the study at any time without penalty or loss of benefits to which you are otherwise entitled. Your decision to take part or not take part will in no way affect your current or future treatment. You will be given ample time and chance to ask questions about the details of this study and to decide if you want to take part.

Once the study has started, if you choose to withdraw, you will need to contact the study doctor. Your reasons for withdrawal will be recorded. For your own safety and interests, you will be asked to undergo an end-of-study visit final exam including a physical exam, blood and urine samples for safety tests, some assessments of any pain you might have, a review of your current medications or treatments. You have the right to refuse to take part in further data collection or follow-up exams after withdrawing from the study. If you withdraw from the study, the sponsor can still use your information that they have already collected.

### 13 Can I be removed from this study without my permission?

The study doctor or the sponsor of this study may decide to withdraw you from the study without your consent. The reasons for withdrawal might be:

- If you are unable to fulfill the requirements of the study.
- If you develop another serious illness.
- If your family doctor believes that continuation of the study is against your interests.
- If the sponsor decides to end the study or your taking part in the study.
- If there is a lack of therapeutic effect, intolerance of the device, unexpected safety events, or any other reason that in the clinical investigator’s opinion, is necessary to protect your safety and welfare.

### 14 What are my responsibilities as a participant in this study?

As a participant in this study you will have to:

- Attend all visits as scheduled.
- Provide honest answers to all questions asked by the study doctor and staff.

The successful conduct of this study depends on the cooperation of study participants. Therefore, we ask that you keep appointments and follow the instructions of your study doctor.

### 15 Who do I contact in case of an emergency?

The study doctor and address of your study site is:

For questions concerning this study or **in case of emergency**, please contact the study doctor at your study site. The Study Doctor will treat you or refer you for treatment. If you are not able to reach the study doctor at the telephone number listed here, please contact your family doctor. If it is a true emergency situation, call 911 or visit the closest emergency department.

### 16 Who do I contact if I have questions about this study?

If you have any questions, concerns, or complaints about the study or your taking part in this study, you may contact:

This research is being overseen by an Institutional Review Board ("IRB"). An IRB is a group of people who perform independent review of research studies. You may talk to them at 855-818-2289 or if:

- You have questions, concerns, or complaints that are not being answered by the research team.
- You are not getting answers from the research team.
- You cannot reach the research team.
- You want to talk to someone else about the research.
- You have questions about your rights as a research subject.

### 17 Will I be informed of new information about this study?

In the event that important new findings become available during the study, which might be relevant to your willingness to continue in the study, this information will be provided to you.

### 18 Who has reviewed this study?

This study will be conducted under the applicable regulatory requirements in the US and Good Clinical Practice rules. These rules are internationally accepted guidelines for performing studies that are safe and produce valid data. The necessary study documents will be submitted to the relevant institutional review board (IRB), which is a group of people who review the ethics of human research. The study will begin only after approval has been received.

### 19 Study available on clinicaltrials.gov website

A description of this clinical trial will be available on http://www.ClinicalTrials.gov, as required by U.S. Law. This Web site will not include information that can identify you. At most, the Web site will include a summary of the results. You can search this Web site at any time.

My taking part in this study is voluntary. I have the right to stop taking part in the study at any time, without specifying my reasons, and without penalties or loss of benefits to which I am otherwise entitled. I understand that it is in my best interest and for the safety of my health to have a final examination and to allow the collection of blood samples necessary to perform laboratory tests for safety reasons. I can also be excluded from further taking part in the study if the study doctor considers it necessary for my safety.

I am ready to follow the instructions given by the study doctor and his or her staff who are conducting the study.

I have answered all questions presented to me to the best of my knowledge.

I confirm with my signature that I have had enough time to read and understand the information in this consent form. My questions have been answered to my satisfaction. I have the right to ask further questions as they come up during the study.

I have been informed of who to contact if anything goes wrong in the study, if I have any questions, or am hurt in any way.

I agree to take part in the study on the basis of the information provided to me by the study doctor conducting the study.

I have received a copy of this form(s).

I agree to the review of my data collected by the study doctor, by persons authorized and obligated to secrecy by the sponsor, the responsible national (and foreign) regulatory authority, or the responsible federal authority as far as this is necessary for the review of the study.

If, during this study, I am hospitalized or receive treatment at a health care facility other than the study site, I agree to allow the study doctor access to medical records related to any treatment received and hospital-release papers.

| | |
|---|---|
| ______________________________ | ______________________________ |
| Signature of the study subject | Date |
| Study subject no.: ____________ | Study subject initials: _____________ |
| ______________________________ | ______________________________ |
| Signature of the investigator | Date |

### Authorization concerning your personal information

For taking part in the research study:

**Pivotal Study of the NanoKnife System for Ablation of Prostate Tissue in an Intermediate Risk Patient Population**

During this study, your personal information and medical findings will be collected. This information will be stored and reviewed according to the law. This requires your consent prior to taking part in the study.

If you choose to be in this study, the study doctor, along with the sponsor AngioDynamics, will gather and use your personal information to conduct the study. The personal information may include the following:

- Your name, address and contact details
- Date of birth
- Demographic information, such as race and gender
- Medical history, including past and present medical records
- Information from your study visits, including test results

The study doctor may share your personal information with:

- AngioDynamics or its designee (any company they use to oversee or conduct the study)
- The IRB, which is a group of people who review the ethics of human research
- A data and safety monitoring board
- The FDA and other US governmental agencies
- The Department of Health and Human Services
- Governmental agencies for other countries
- Other companies or agencies working with (or owned by) the sponsor and its designees
- Your family doctor, as necessary

The sponsor and those working for them may use the personal information sent to them:

- To see if the study device works
- To see if the study device is safe

- For other research activities related to the study device
- To ensure that applicable laws and procedures are being followed by the study
- To make any reports required by applicable laws

Once your personal information has been shared with authorized users, it may no longer be protected by privacy laws.

Your personal information will be retained for a period determined by applicable laws and regulatory requirements, which could be for the entire life of the study device or a longer period.

The consent you provide today. You may withdraw your consent to use and share your personal information at any time by notifying the study doctor, preferably in writing. You can do this by calling or sending written notice to the study doctor at the address in **Section 16**. If you withdraw your consent, you will not be able to stay in this study. When you withdraw your consent, no new personal information about you will be collected after that date. However, the personal information that has already been collected may still be used and given to others.

You do not have to sign this form but please be aware, that if you decide not to sign this form, you will not be able to be in the study. Your decision to not sign this form will not affect any of your other treatment, health care, enrollment in health plans or eligibility for benefits.

### Authorization

I have read this form and its contents were explained to me. I have had a chance to ask questions, and my questions have been answered. I voluntarily agree to allow the study staff to collect, use, and share my personal information in the manner and for the purposes outlined above. I will receive a signed copy of this form for my records.

| | |
|---|---|
| ______________________ | ______________________ |
| Signature of the study subject | Date |
| Study subject no.: __________ | Study subject initials: ___________ |
| ______________________ | ______________________ |
| Signature of the investigator | Date |
